# Association of regional Covid-19 mortality with indicators of indoor ventilation, including temperature and wind: insights into the upcoming winter

**DOI:** 10.1101/2021.12.05.21267334

**Authors:** Christopher T. Leffler, Joseph D. Lykins V, Brandon I. Fram, Edward Yang

## Abstract

**Background:** Outdoor environmental variables, such as cold temperatures and low wind speed, have been correlated with incidence and mortality from Covid-19 (caused by the SARS-CoV-2 virus). However, as Covid-19 predominantly spreads indoors, the degree to which outdoor environmental variables might directly cause disease spread is unclear.

**Methods:** World regions were considered to have reliable data if the excess mortality did not greatly exceed reported Covid-19 mortality. The relative risk of Covid-19 mortality for 142 regions as a function of median weekly temperature and wind speed was determined. For instance, Covid-19 mortality following warm weeks in a country was compared with mortality following cold weeks in the same country.

**Results:** Covid-19 mortality increases with cooling from 20 C to close to freezing (0 to 4 C, p<0.001). The relation of Covid-19 mortality with temperature demonstrates a maximum close to freezing. Below -5 C, the decrease in mortality with further cooling was statistically significant (p<0.01). With warming above room temperature (20 to 24 C), there is a nonsignificant trend for mortality to increase again. A literature review demonstrated that window opening and indoor ventilation tend to increase with warming in the range from freezing to room temperature.

**Conclusion:** The steep decline in Covid-19 mortality with warming in the range from freezing to room temperature may relate to window opening and less indoor crowding when it is comfortable outside. Below freezing, all windows are closed, and further cooling increases stack ventilation (secondary to indoor-outdoor temperature differences) and thereby tends to decrease Covid-19 mortality. Opening windows and other tools for improving indoor ventilation may decrease the spread of Covid-19.

## Introduction

The Covid-19 pandemic (caused by the SARS-CoV-2 virus) has spread throughout the world, often in multiple waves in individual countries. The timing of these waves of infection has been difficult for public health authorities to predict.

The association of outdoor environmental variables, such as temperature, humidity, and wind speed, with the spread of Covid-19 has been studied. One primary finding is that colder temperatures are associated with a higher incidence of Covid-19 infection and mortality (Wu 2020).

However, there are both empirical and logical reasons to question the degree to which these outdoor environmental variables might directly cause the spread of Covid-19. First, the empirical findings. Some investigators have found that Covid-19 is positively (rather than negatively) associated with temperature below 3 C (37.4° F) (Xie 2020). Others have found that Covid-19 incidence peaks between 5 C and 15 C (rather than continuing to rise at the coldest temperatures) (Huang 2020). In addition, a peak in Covid-19 incidence was observed in the sunbelt of the United States in the summer of 2020, and in the warmer months of April and May of 2021 in India (Leffler 2021).

On logical grounds, it has been unclear why outdoor environmental variables might be causatively linked with Covid-19 in a direct fashion, given that Covid-19 is believed to primarily spread indoors.

However, we noted that there is a large body of literature demonstrating that the major predictor of indoor ventilation is in fact outdoor temperature, with wind speed generally playing a lesser role. The reason is that temperature influences both occupant window-opening behavior and stack ventilation resulting from indoor-outdoor temperature differences.

We therefore sought to analyze the association of mortality from Covid-19 with environmental variables, from the perspective of their potential impact on indoor ventilation.

## Methods

We selected countries for analysis if the ratio of excess mortality during the pandemic to Covid-19 mortality from the World Mortality Dataset did not exceed 1.8 (Karlinsky 2021, Kobak 2021). Our reasoning was that if the Covid-19 mortality was well below the excess mortality, the country was probably under-reporting Covid-19 mortality, either due to poor testing levels or suppression of information. This threshold was admittedly arbitrary, but it did serve to objectively include countries reputed to have developed and open health systems (such as Finland, with a ratio of 1.7), while excluding countries believed to have suppressed information about the pandemic, such as Iran (ratio 2.0) and Russia (ratio 3.6). This method also excluded countries with more closed health systems, such as China, for which no excess mortality data were available in the World Mortality Dataset.

Covid-19 mortality data for each week of the pandemic were downloaded from the Johns Hopkins University Center for Systems Science and Engineering on October 3, 2021 (Hopkins 2021). For some countries, particularly smaller countries, this dataset merely lists Covid-19 mortality for the entire country. For other countries, such as the United States, Canada, Brazil, and Chile, the dataset lists mortality for states or provinces separately. Among the countries analyzed, we selected states (or provinces) for analysis by listing the most populous city in each of the country’s states, and starting with the most populous such city, accepting the city and state for analysis if the state had at least 5 reported Covid-19 deaths, and if the city was at least 500 km from another of the country’s cities accepted for analysis. Thus, for states close to each other, such as New York and New Jersey, only the former was accepted into the analysis. The reason to exclude cities within 500 km of each other is that cities close to each other could be subject to essentially the same weather system.

For some countries, the Hopkins dataset contains a week with small decreases in cumulative mortality for the week, which would appear to represent negative deaths for the week. These anomalies were handled by eliminating the week with apparent negative deaths. In the case of Belgium, the worldometer data were used, because the data appeared similar, but did not have this anomaly.

For each accepted city, the daily temperature, dew point, and wind speed during the pandemic were downloaded from the National Oceanic and Atmospheric Administration (NOAA 2021). The median weekly temperature (median of all daily maxima and minima) was calculated.

It is on average 23 days from exposure to Sars-CoV-2 and death (Leffler 2020). Therefore, we analyzed the association of median weekly temperature with the weekly mortality 3 weeks later. In essence, the time between each exposure day and each outcome day was between 14 and 28 days, which straddled the 23 day average lag between exposure and death. For each exposure week, the relative risk of Covid-19 mortality was determined. The relative risk was calculated as observed Covid-19 deaths for the week divided by expected Covid-19 deaths. The expected deaths were defined as the total number of the region’s Covid-19 deaths in the dataset divided by the number of weeks of the pandemic with data for that region. The pandemic start for a region was defined as the first week in the dataset with a Covid-19 death, or the fourth week in the dataset after Covid-19 cases were recorded in the region in the dataset (whichever came first).

The relative risk of Covid-19 at each temperature was depicted graphically. For each temperature within one country’s data, the average relative risk for that country at that temperature was obtained. Then, the relative risks for all countries at that temperature were averaged. Thus, each country contributed to the analysis at each temperature just one time, at most. For temperatures of -8 C (−17.6° F) or below, or 33 C (91.4° F) or above, there were fewer than 10 observations. Therefore, in order to obtain reliable estimates at these extreme temperatures, data from neighboring temperatures were included until at least 10 observations were available, and the mean relative risk was plotted against the median temperature in the observation set.

The household prevalence of air-conditioning in each country was obtained from reports of the International Energy Agency (IEA 2018, 2019), which published either the actual prevalence, or the number of air-conditioning units per household. A report on sales volume was used to calculate the number of air-conditioning units sold annually per capita in multiple countries (Japan 2019). The IEA reports were used to determine the relationship between prevalence and per-capita sales. Once this relationship was determined, then sales volume in other countries could be used to determine household prevalence. Finally, the dependence of air-conditioning prevalence on gross domestic productivity per capita and mean annual temperature was used to estimate the prevalence in countries not covered by the previous reports. The estimation of regional household air-conditioning prevalence will be covered in detail in an appendix.

### Statistical analysis

Pairs of temperature ranges were compared by identifying regions which had at least one death within each of the two ranges. The risk ratio for each country was calculated as the ratio of weekly mortality in the two temperature ranges. The logarithm of the risk ratio was compared with zero by a one-sample t-test. The regional mortality data studied contained no individually identifiable information. The study was approved by the Office of Research Subjects Protection at Virginia Commonwealth University.

## Results

### Temperature

We identified 142 regions in 68 countries for analysis (Table 1). The graph of Covid-19 mortality as a function of outdoor temperature had two distinct peaks, separated by a local minimum at room temperature (20 C, 68° F) (Figure 1, Tables 2 to 4).

**Table 1.** World regions analyzed for Covid-19 mortality and weather.

| **Country.** | **City for Weather Data.** | **Region for Covid-19 data** |
| --- | --- | --- |
| Antigua and Barbuda | Saint Johns | Antigua and Barbuda |
| Argentina | Buenos Aires | Argentina |
| Aruba | Oranjestad | Aruba |
| Australia | Brisbane | Southern Australia |
| Australia | Hobart | Tasmania |
| Australia | Melbourne | Victoria |
| Australia | Perth | Western Australia |
| Australia | Sydney | New South Wales |
| Austria | Vienna | Austria |
| Belgium | Antwerp | Belgium |
| Belize | Belize City | Belize |
| Bosnia | Sarajevo | Bosnia |
| Brazil | Belem | Para |
| Brazil | Boa Vista | Roraima |
| Brazil | Brasilia | Distrito Federal |
| Brazil | Campo Grande | Mato Grosso do sul |
| Brazil | Cuiabá | Mato Grosso |
| Brazil | Fortaleza | Ceara |
| Brazil | Manaus | Amazonas |
| Brazil | Palmas | Tocantins |
| Brazil | Porto Alegre | Rio Grande do Sul |
| Brazil | Porto Velho | Rondonia |
| Brazil | Recife | Pernambuco |
| Brazil | Salvador | Bahia |
| Brazil | Sao Paulo | Sao Paulo |
| Canada | Calgary | Alberta |
| Canada | Halifax | Nova Scotia |
| Canada | Montreal | Quebec |
| Canada | Saskatoon | Saskatchewan |
| Canada | St. Johns | Newfoundland and Labrador |
| Canada | Toronto | Ontario |
| Canada | Vancouver | British Columbia |
| Canada | Whitehorse | Yukon |
| Canada | Winnipeg | Manitoba |
| Canada | Yellowknife | Northwest Territories |
| Chile | Antofagasta | Antofagasta |
| Chile | Arica | Arica y Parinacota |
| Chile | Coyhaique | Aysen |
| Chile | Punta Arenas | Magallanes |
| Chile | Santiago | Metropolitana |
| Chile | Temuco | Araucania |
| Colombia | Barranquilla | Atlántico |
| Colombia | Bogota | Capital District |
| Colombia | Leticia | Amazonas |
| Costa Rica | San Jose | Costa Rica |
| Croatia | Zagreb | Croatia |
| Cyprus | Nicosia | Cyprus |
| Czechia | Prague | Czechia |
| Denmark | Copenhagen | Denmark |
| Estonia | Tallinn | Estonia |
| Finland | Helsinki | Finland |
| France | Cayenne | French Guiana |
| France | Fort-de-France | Martinique |
| France | Les Abymes | Guadeloupe |
| France | Paris | France, continental |
| France | Saint-Denis | Reunion |
| French Polynesia | Faaa | French Polynesia |
| Georgia | Tbilisi | Georgia |
| Germany | Berlin | Berlin |
| Germany | Munich | Bavaria |
| Gibraltar | Gibraltar | Gibraltar |
| Greece | Athens | Greece |
| Hungary | Budapest | Hungary |
| Iceland | Reykjavík | Iceland |
| Ireland | Dublin | Ireland |
| Israel | Tel Aviv | Israel |
| Italy | Milan | Lombardy |
| Italy | Reggio Calabria | Calabria |
| Italy | Rome | Lazio |
| Jamaica | Kingston | Jamaica |
| Japan | Fukuoka | Fukuoka prefecture |
| Japan | Naha | Okinawa |
| Japan | Sapporo | Hokkaido |
| Japan | Tokyo | Tokyo |
| Kosovo | Prishtina | Kosovo |
| Latvia | Riga | Latvia |
| Lebanon | Beirut | Lebanon |
| Liechtenstein | Schaan | Liechtenstein |
| Luxembourg | Luxembourg | Luxembourg |
| Malaysia | Kota Kinabalu | Sabah |
| Malaysia | Kuala Lumpur | Selangor |
| Malaysia | Kuching | Sarawak |
| Malta | Birkirkara | Malta |
| Mauritius | Port Louis | Mauritius |
| Moldova | Chisinau | Moldova |
| Mongolia | Ulaanbaatar | Mongolia |
| Montenegro | Podgorica | Montenegro |
| Netherlands | Amsterdam | Netherlands |
| New Zealand | Auckland | New Zealand |
| Norway | Oslo | Norway |
| Oman | Muscat | Oman |
| Panama | Panama City | Panama |
| Paraguay | Asuncion | Paraguay |
| Peru | Arequipa | Arequipa |
| Peru | Chiclayo | Lambayeque |
| Peru | Iquitos | Loreto |
| Peru | Lima | Lima |
| Peru | Pucallpa | Ucayali |
| Poland | Warsaw | Poland |
| Portugal | Lisbon | Portugal |
| Qatar | Doha | Qatar |
| Romania | Bucharest | Romania |
| San Marino | San Marino | San Marino |
| Seychelles | Victoria | Seychelles |
| Slovakia | Bratislava | Slovakia |
| Slovenia | Ljubljana | Slovenia |
| South Korea | Seoul | South Korea |
| Spain | A Coruña–Oleiros–Arteixo | Galicia |
| Spain | Barcelona | Catalonia |
| Spain | Madrid | Madrid |
| Sweden | Malmo | Skane |
| Sweden | Stockholm | Stockholm |
| Sweden | Umea | Vasterbotten County |
| Switzerland | Zurich | Switzerland |
| Taiwan | Taipei | Taiwan |
| Tunisia | Tunis | Tunisia |
| Ukraine | Kiev | Kiev |
| Ukraine | Mariupol | Donetsk Oblast |
| Ukraine | Uzhhorod | Zakarpattia Oblast |
| United Kingdom | Glasgow | Scotland |
| United Kingdom | London | England |
| United States | Albuquerque | Bernalillo County |
| United States | Anchorage | Anchorage |
| United States | Billings, Montana | Yellowstone county, Montana |
| United States | Boise, Idaho | Ada County |
| United States | Charlotte | Mecklenburg County |
| United States | Chicago | Cook County, Illinois |
| United States | Cleveland | Cuyahoga County |
| United States | Denver | Denver |
| United States | Honolulu | Honolulu |
| United States | Houston | Houston plus Harris County |
| United States | Jacksonville | Duval County |
| United States | Kansas City, MO | Kansas City, MO |
| United States | Los Angeles | Los Angeles |
| United States | Minneapolis | Hennepin County |
| United States | Nashville | Davidson County |
| United States | New Orleans | Orleans Parish |
| United States | New York City | New York City |
| United States | Oklahoma City | Oklahoma City |
| United States | Phoenix | Maricopa County |
| United States | Seattle | King County |
| Uruguay | Montevideo | Uruguay |

**Figure 1.**
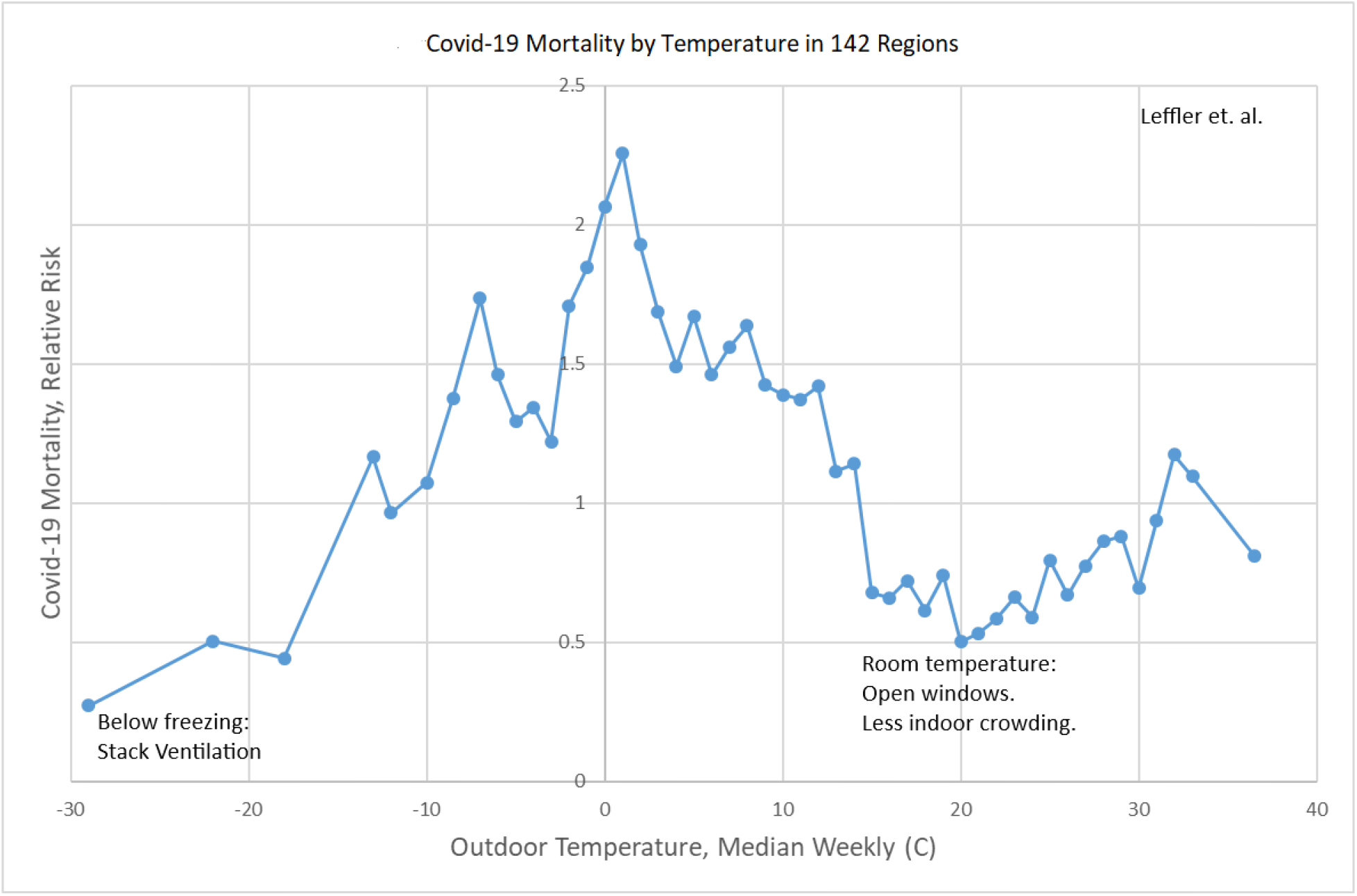
Weekly Covid-19 mortality as a function of median weekly outdoor temperature 3 weeks previously in 142 regions.

**Table 2.** Comparison with mortality close to room temperature (20 to 24 C).

| Temperature | n | Risk ratio (95% CI) | P |
| --- | --- | --- | --- |
| <= -11 C | 12 | 3.46 (1.88 to 6.36) | 0.002 |
| -10 to -6 C | 19 | 8.13 (4.97 to 13.31) | <0.001 |
| -5 to -1 C | 39 | 7.25 (4.86 to 10.82) | <0.001 |
| 0 to 4 C | 59 | 6.84 (4.98 to 9.40) | <0.001 |
| 5 to 9 C | 71 | 5.32 (3.85 to 7.36) | <0.001 |
| 10 to 14 C | 89 | 3.59 (2.76 to 4.66) | <0.001 |
| 15 to 19 C | 98 | 1.58 (1.33 to 1.88) | <0.001 |
| 20 to 24 C | -- | 1.00 | -- |
| 25 to 29 C | 82 | 1.04 (0.85 to 1.26) | 0.71 |
| ≥ 30 C | 31 | 1.23 (1.97 to 0.97) | 0.39 |

n = number of regions.

**Table 3.** Comparison with mortality close to freezing (0 to 4 C).

| Temperature | n | Risk ratio (95% CI) | P |
| --- | --- | --- | --- |
| <= -11 C | 15 | 0.56 (0.30 to 1.04) | 0.09 |
| -10 to -6 C | 22 | 0.88 (0.59 to 1.30) | 0.52 |
| -5 to -1 C | 44 | 0.90 (0.76 to 1.06) | 0.21 |
| 0 to 4 C | -- | 1.00 | -- |
| 5 to 9 C | 68 | 0.81 (0.70 to 0.94) | 0.007 |

n = number of regions.

**Table 4.** Comparison of mortality with cooling through successive temperature ranges.

| Temperature | Reference | N | Risk Ratio (95% CI) | P |
| --- | --- | --- | --- | --- |
| <= -11 C | -10 to -6 C | 12 | 0.57 (0.41 to 0.80) | 0.007 |
| -10 to -6 C | -5 to -1 C | 21 | 0.38 (0.21 to 0.67) | 0.004 |
| -5 to -1 C | 0 to 4 C | 44 | 0.90 (0.76 to 1.06) | 0.21 |
| 0 to 4 C | 5 to 9 C | 68 | 1.24 (1.06 to 1.44) | 0.007 |
| 5 to 9 C | 10 to 14 C | 81 | 1.44 (1.23 to 1.68) | <0.001 |
| 10 to 14 C | 15 to 19 C | 96 | 2.01 (1.69 to 1.40) | <0.001 |
| 15 to 19 C | 20 to 24 C | 98 | 1.58 (1.33 to 1.88) | <0.001 |
| 20 to 24 C | 25 to 29 C | 82 | 0.96 (0.87 to 1.17) | 0.71 |
| 25 to 29 C | ≥ 30 C | 41 | 0.74 (0.63 to 1.02) | 0.07 |

n = number of regions.

### Air-Conditioning

To determine if air-conditioning might influence the dependence of Covid-19 mortality on temperature, we analyzed separately regions estimated to have a low household prevalence of air-conditioning (<25%) compared with those with a higher prevalence (>50%, Figure 2, Table 5). The graphical profiles do not look very different between the two groups, though the data are noisy (Figure 2). Interestingly, the elevation in relative risk with cooling to 14 C (57.2° F) is more abrupt in the regions with more air-conditioning.

**Figure 2.**
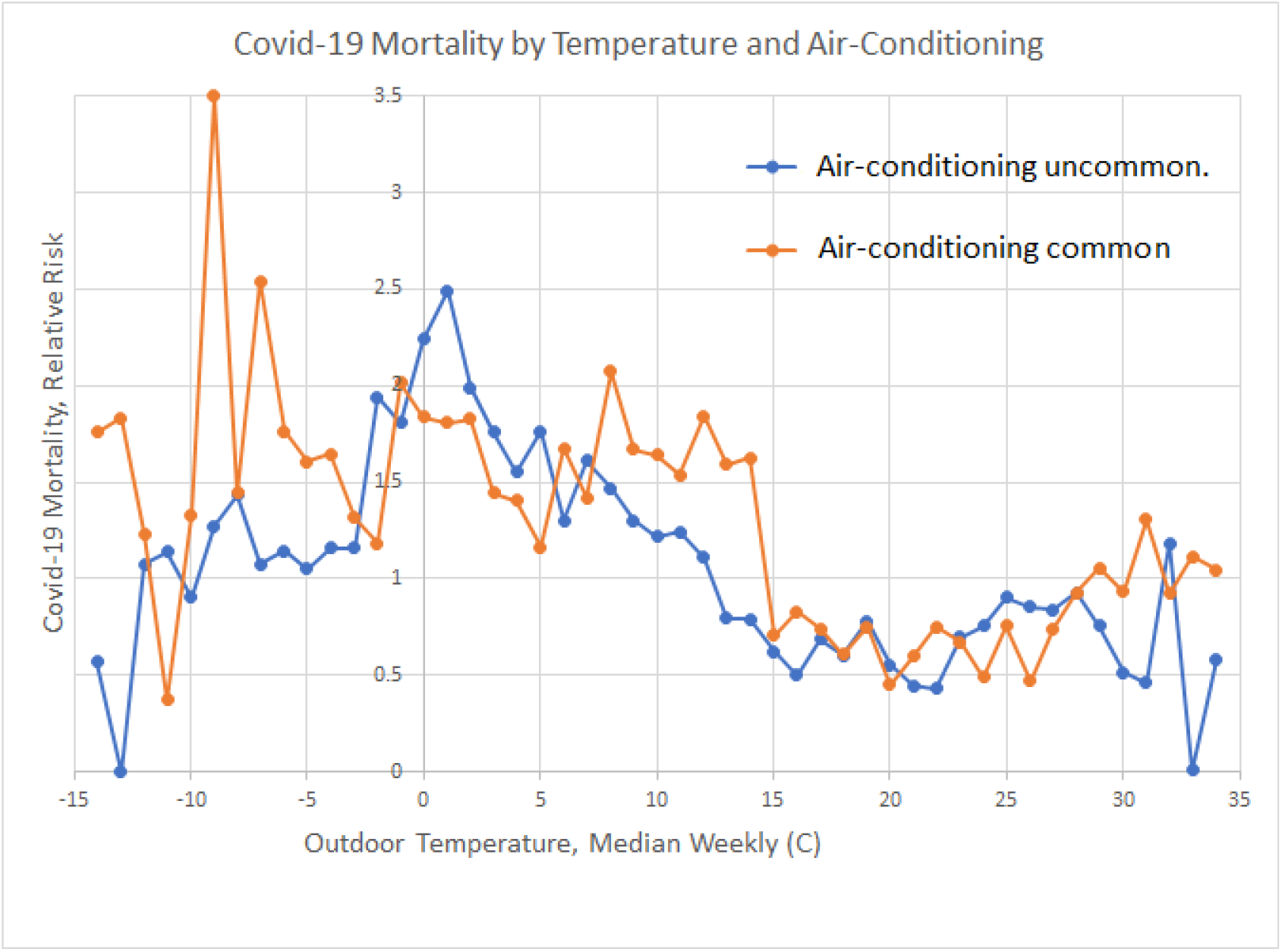
Weekly Covid-19 mortality as a function of median weekly outdoor temperature 3 weeks previously, by whether air conditioning prevalence was uncommon (<25%), or common (>=50%).

**Table 5.** Regions with low and high prevalence of air-conditioning.

| Air-conditioning Prevalence | Regions. |
| --- | --- |
| <25% | Argentina, Austria, Belgium, Belize, Bosnia and Herzegovina, Brazil, Canada (British Columbia, Newfoundland and Labrador, Northwest Territories, Nova Scotia, Yukon), Chile, Colombia, Costa Rica, Croatia, Czechia, Denmark, Estonia, Finland, France, Georgia, Hungary, Iceland, Ireland, Jamaica, Kosovo, Latvia, Lebanon, Liechtenstein, Luxembourg, Moldova, Mongolia, Montenegro, New Zealand, Netherlands, Norway, Panama, Paraguay, Peru, Poland, Portugal, Romania, Slovakia, Slovenia, Sweden, Switzerland, Tunisia, Ukraine, United Kingdom, United States (Alaska only), Uruguay. |
| ≥50% | Aruba, Australia, Canada (Manitoba, Ontario, Saskatchewan), Gibraltar, Greece, Guadeloupe, Israel, Japan, Malaysia, Malta, Martinique, Oman, Qatar, Reunion, San Marino, South Korea, Spain, Taiwan, United States (except Alaska and Washington state). |

### Wind Speed

Next, we analyzed the association of weekly Covid-19 mortality with the median weekly wind speed 3 weeks previously. The relative risk of Covid-19 appears to be elevated with the lowest wind speeds (below 3 m/s). At or above 3 m/s, the relative risk is somewhat lower, though above 11 m/s, the data are noisy because of fewer available observations.

But, given that Covid-19 typically spreads indoors, we might consider the effect of the impact of wind speed on indoor ventilation. Windows are more likely to be closed when it is cold outside, especially below 15 C (59° F). Therefore, if one assumes the impact of the wind on indoor ventilation is zero with a temperature of 15 C (59° F) or below, then wind speed can be replaced with a value of zero with these cold temperatures. The resulting association does point to lower Covid-19 mortality with higher wind speeds, but again with some variation in the data (Figure 4).

**Figure 3.**
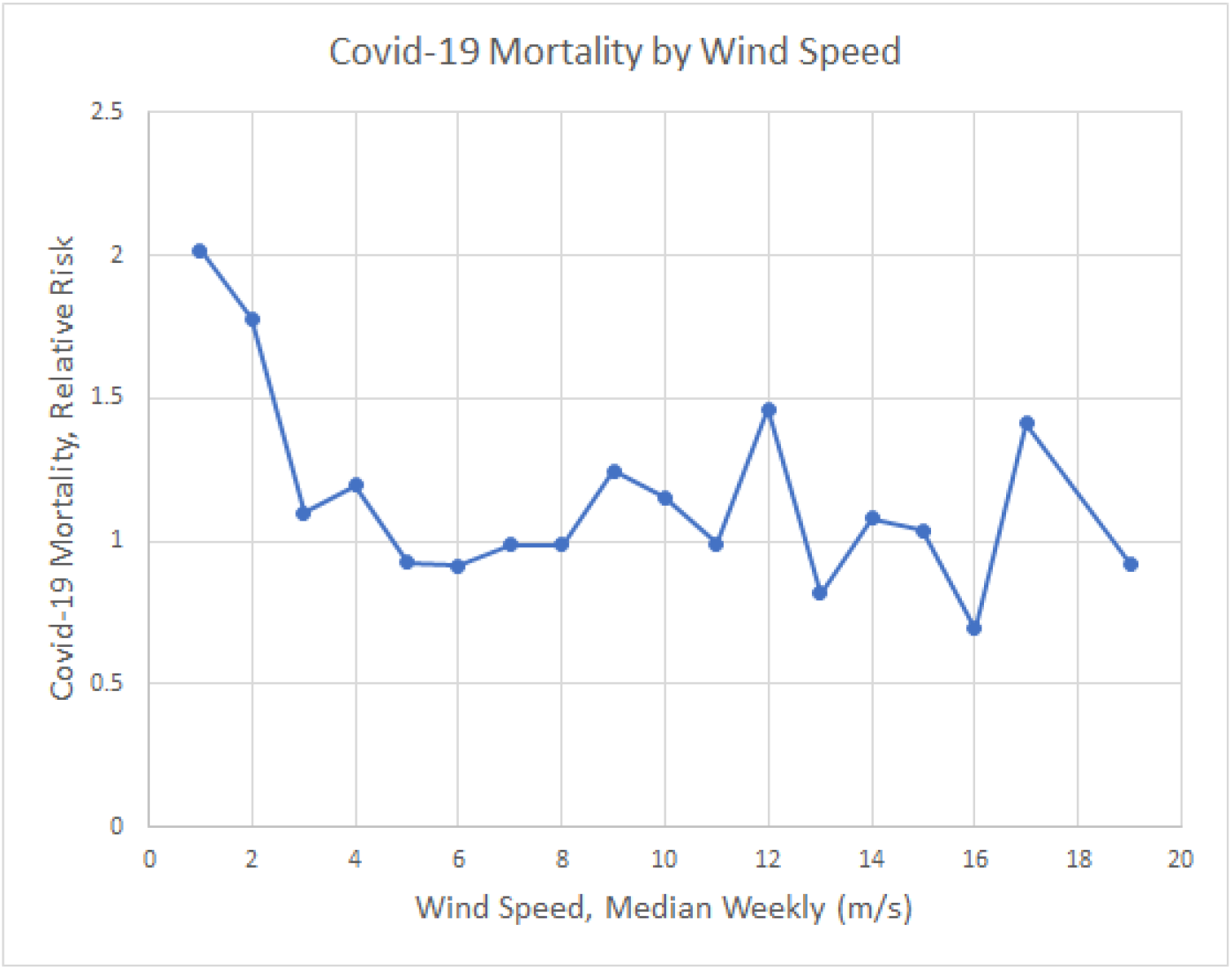
Weekly Covid-19 mortality as a function of median weekly wind speed 3 weeks previously.

**Figure 4.**
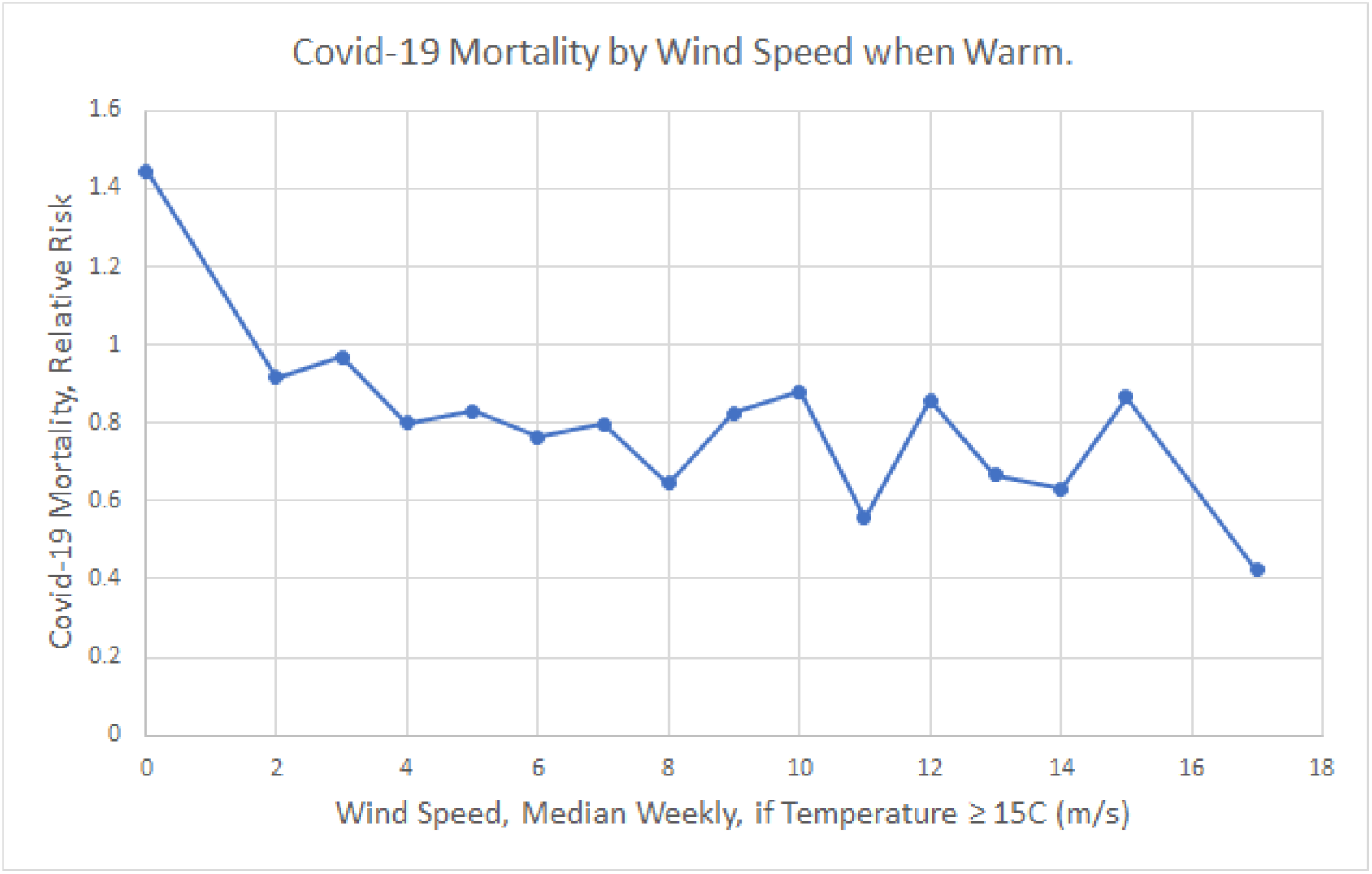
Weekly Covid-19 mortality as a function of median weekly wind speed 3 weeks previously, assuming that due to window closure, effective wind speed able to be transmitted indoors is zero when outdoor temperature is at or below 15 C (59° F).

We also plotted the relationship of Covid-19 mortality as a function of wind speed, in which the effective wind speed transmitted indoors was also estimated to be zero due to window closure in predominantly air-conditioned regions when the outdoor temperature was over 30 C (86° F). The plot was essentially unchanged, and is therefore not included.

Next, we analyzed the relationship between weekly Covid-19 mortality and median weekly outdoor dew point 3 weeks previously (Figure 5). The two-peaked curve is similar to the relationship of mortality with outdoor temperature, but the peaks and valleys are flatter. Whether humidity provides information on mortality independent of temperature must await multivariable analysis.

**Figure 5.**
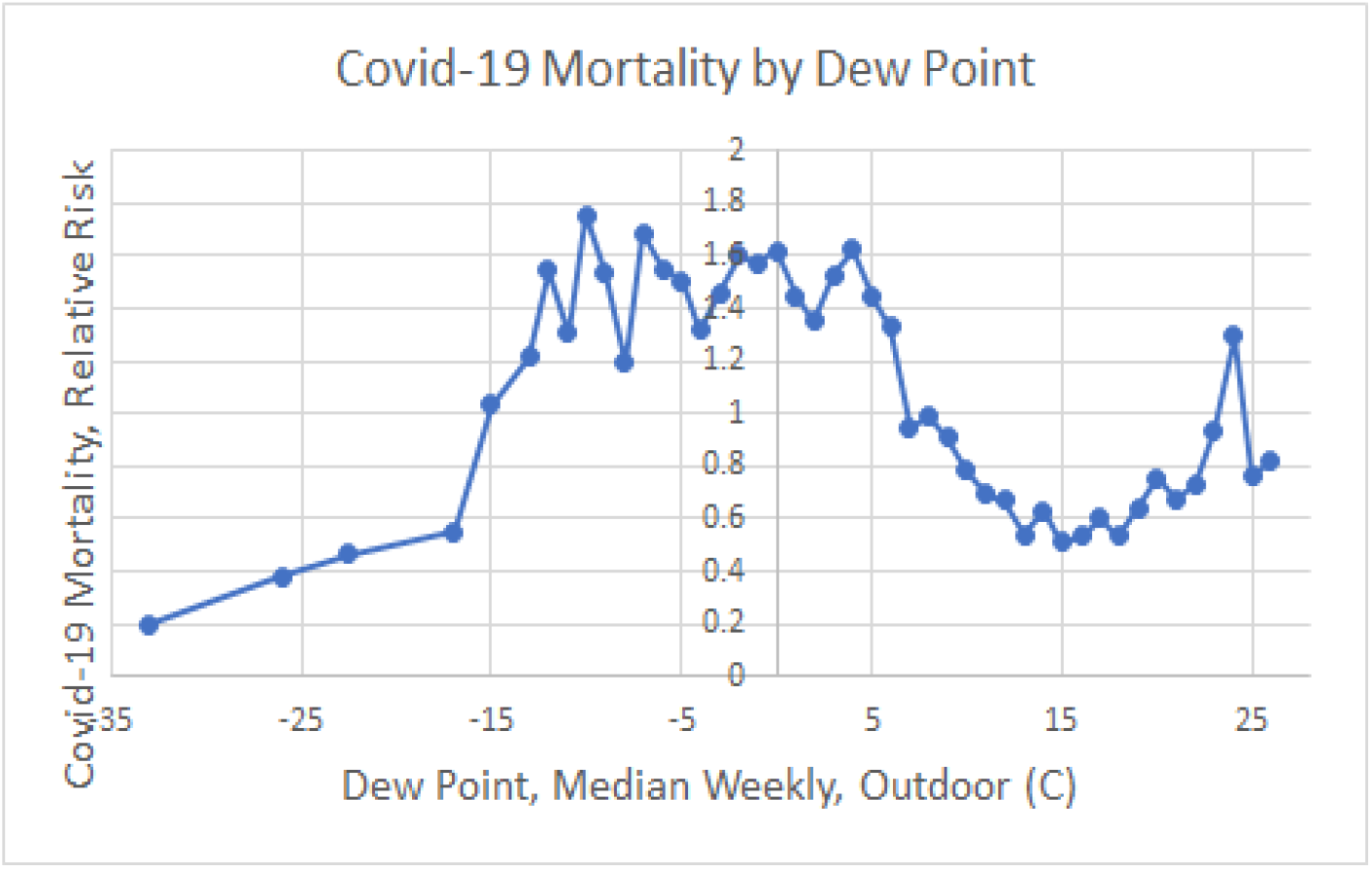
Weekly Covid-19 mortality as a function of median weekly outdoor dew point 3 weeks previously.

### Window-opening and outdoor temperature

A literature review demonstrated that outdoor temperature was a major determinant of occupant window-opening behavior in both homes and workplaces (Table 6, Figure 6).

**Table 6.**
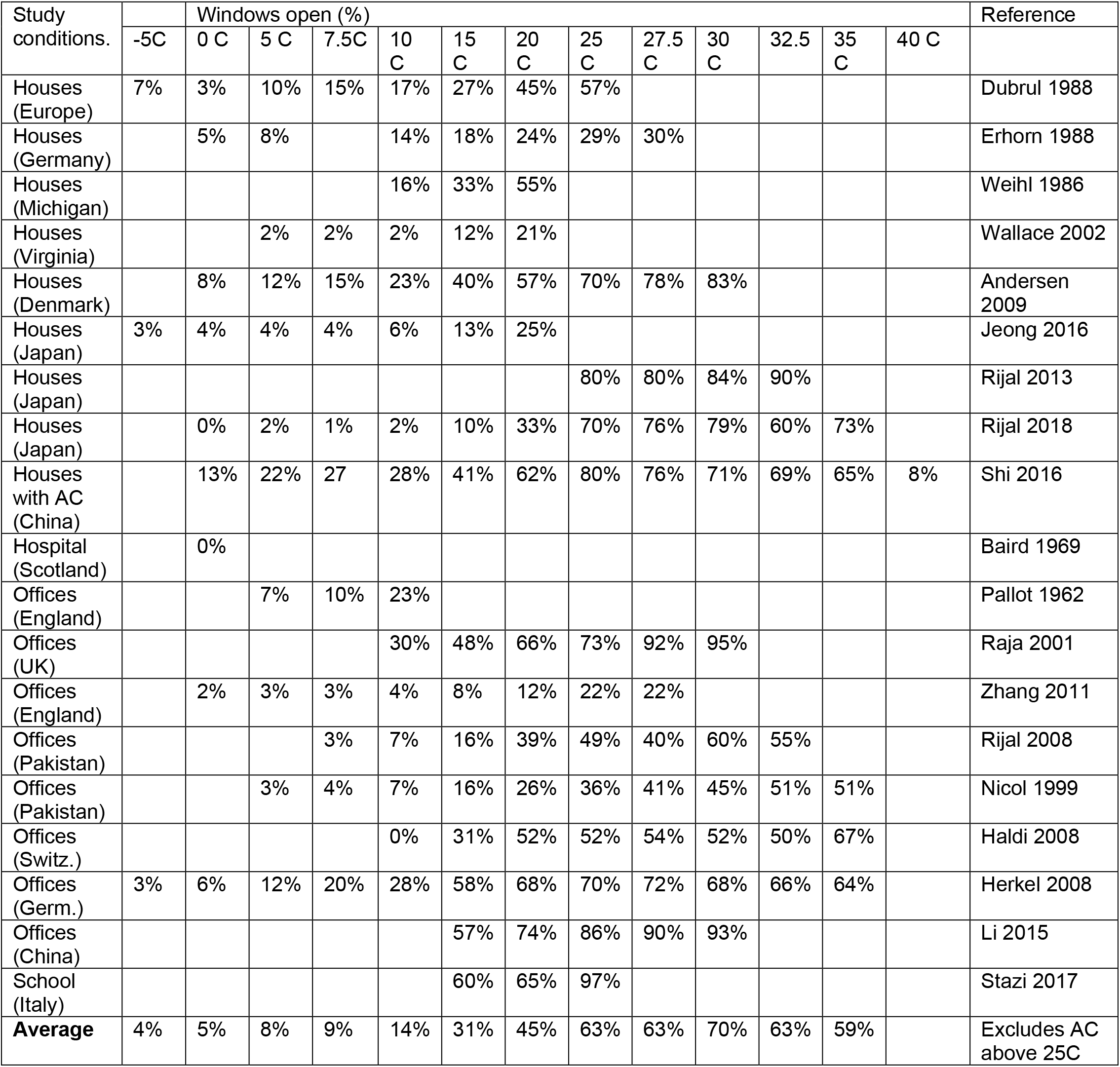
Association of window-opening with outdoor temperature.

| Study conditions. | Windows open (%) |  |  |  |  |  |  |  |  |  |  |  |  | Reference |
| --- | --- | --- | --- | --- | --- | --- | --- | --- | --- | --- | --- | --- | --- | --- |
|  | -5C | 0 C | 5 C | 7.5C | 10 C | 15 C | 20 C | 25 C | 27.5 C | 30 C | 32.5 | 35 C | 40 C |  |
| Houses (Europe) | 7% | 3% | 10% | 15% | 17% | 27% | 45% | 57% |  |  |  |  |  | Dubrul 1988 |
| Houses (Germany) |  | 5% | 8% |  | 14% | 18% | 24% | 29% | 30% |  |  |  |  | Erhorn 1988 |
| Houses (Michigan) |  |  |  |  | 16% | 33% | 55% |  |  |  |  |  |  | Weihl 1986 |
| Houses (Virginia) |  |  | 2% | 2% | 2% | 12% | 21% |  |  |  |  |  |  | Wallace 2002 |
| Houses (Denmark) |  | 8% | 12% | 15% | 23% | 40% | 57% | 70% | 78% | 83% |  |  |  | Andersen 2009 |
| Houses (Japan) | 3% | 4% | 4% | 4% | 6% | 13% | 25% |  |  |  |  |  |  | Jeong 2016 |
| Houses (Japan) |  |  |  |  |  |  |  | 80% | 80% | 84% | 90% |  |  | Rijal 2013 |
| Houses (Japan) |  | 0% | 2% | 1% | 2% | 10% | 33% | 70% | 76% | 79% | 60% | 73% |  | Rijal 2018 |
| Houses with AC (China) |  | 13% | 22% | 27 | 28% | 41% | 62% | 80% | 76% | 71% | 69% | 65% | 8% | Shi 2016 |
| Hospital (Scotland) |  | 0% |  |  |  |  |  |  |  |  |  |  |  | Baird 1969 |
| Offices (England) |  |  | 7% | 10% | 23% |  |  |  |  |  |  |  |  | Pallot 1962 |
| Offices (UK) |  |  |  |  | 30% | 48% | 66% | 73% | 92% | 95% |  |  |  | Raja 2001 |
| Offices (England) |  | 2% | 3% | 3% | 4% | 8% | 12% | 22% | 22% |  |  |  |  | Zhang 2011 |
| Offices (Pakistan) |  |  |  | 3% | 7% | 16% | 39% | 49% | 40% | 60% | 55% |  |  | Rijal 2008 |
| Offices (Pakistan) |  |  | 3% | 4% | 7% | 16% | 26% | 36% | 41% | 45% | 51% | 51% |  | Nicol 1999 |
| Offices (Switz.) |  |  |  |  | 0% | 31% | 52% | 52% | 54% | 52% | 50% | 67% |  | Haldi 2008 |
| Offices (Germ.) | 3% | 6% | 12% | 20% | 28% | 58% | 68% | 70% | 72% | 68% | 66% | 64% |  | Herkel 2008 |
| Offices (China) |  |  |  |  |  | 57% | 74% | 86% | 90% | 93% |  |  |  | Li 2015 |
| School (Italy) |  |  |  |  |  | 60% | 65% | 97% |  |  |  |  |  | Stazi 2017 |
| **Average** | 4% | 5% | 8% | 9% | 14% | 31% | 45% | 63% | 63% | 70% | 63% | 59% |  | Excludes AC above 25C |

**Figure 6.**
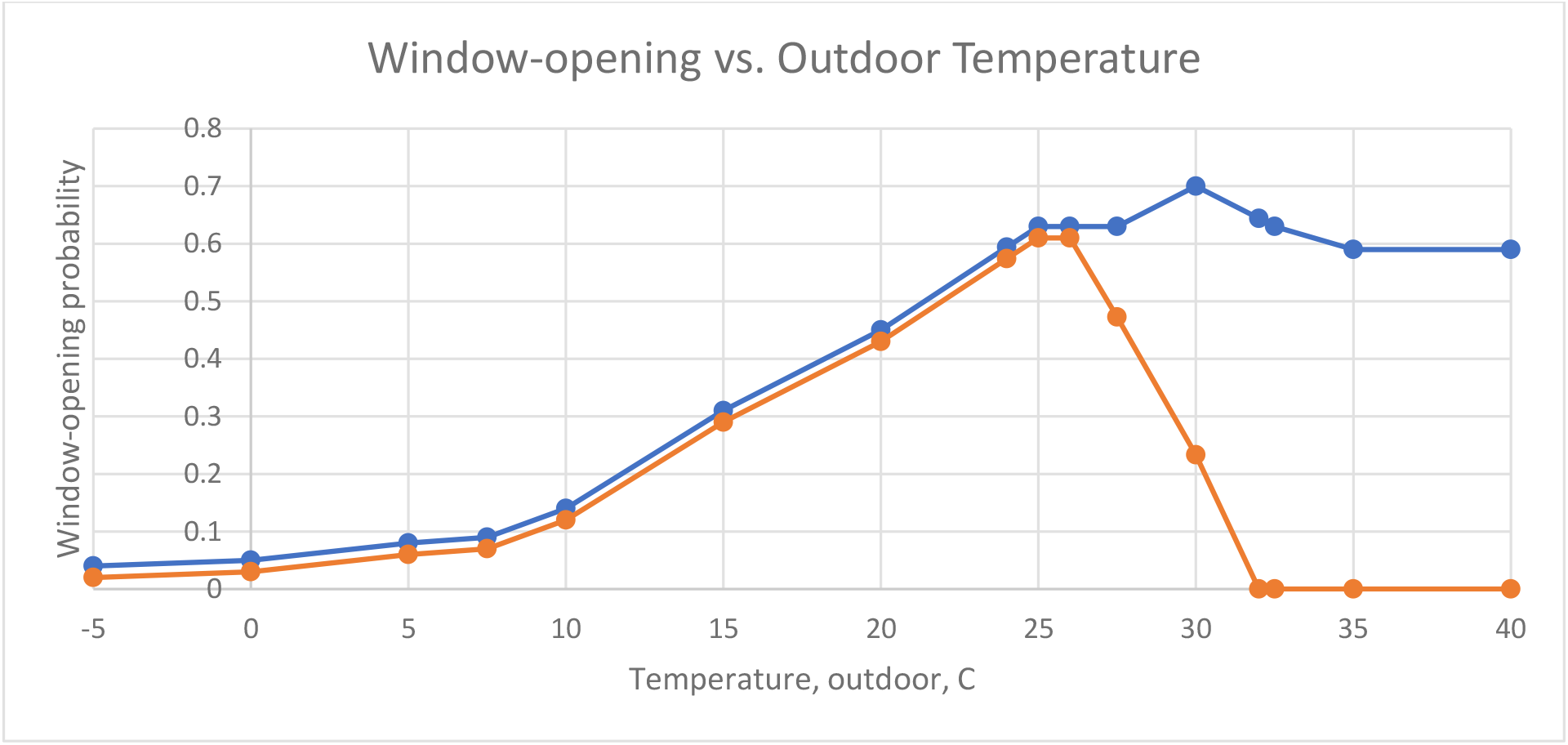
The probability of window-opening as a function of outdoor temperature, without air conditioning (blue line), and in households with air conditioning (orange line), based on the figures from the literature in Table 3.

Traditionally, people close windows when it is very cold outside to keep the building from getting too cold. Between freezing and room temperature, the number of windows (Dick 1951, Brundrett 1977, Brundrett 1978), area of window opening (Baird 1969), and the probability of window-opening (Pallot 1962; Dubrul 1988; Erhorn 1988) are roughly linear. The probability of window-opening progressively increases, reaching a maximum close to or just above room temperature in some studies (Rijal 2008; Rijal 2013; Herkel 2008). Window-opening increases progressively with momentary (Pallot 1962), mean monthly (Pallot 1962; Wallace 2002), and seasonal (Weihl 1990) temperature.

Windows in German homes were typically open a minimum of 5% of the time in February (mean 3 C) and a maximum of 27% of the time in August (mean 20 C; Erhorn 1988).

Window-opening in an office building increased from 5% in December (mean 36 F) to 23% in April and October (mean 47-51 °F; Pallot 1962). Windows were open <2% of the time at 30 °F all year (Pallot 1962). When it was 50 °F outside, windows were open just 11% of the time in December, 21% of the time in October, and 29% of the time in April (Pallot 1962). Thus, people tend to close the windows more in the colder seasons, even when it happens to be warm outside (Pallot 1962).

In an English school, window-opening was at a minimum of 14% in January, and a maximum of 85% in September (Davies 1987). (No August data were available.)

In Michigan in 1986, homes without air conditioning tended to have windows open 2 min per hour in Winter (29.4 °F), 14 min per hour in Spring (53.1 °F), 33 min per hour in Summer (68.6 °F), and 7 min per hour in the Fall (51.2 °F) (Weihl 1986).

Traditionally, people have controlled the ventilation in their homes and workplaces by opening windows. As anyone who has burned something in the kitchen knows, the smoke can be cleared out much more quickly if windows are opened, as the ventilation increases. Indeed, studies show that window-opening can substantially improve indoor ventilation (Andersen 2009). The average household air-exchange rate can increase from 1.75 per hour at 33 °F to 3.25 per hour at 48 °F, largely due to window-opening behavior (Dick 1951).

With a wind speed of 2-4 m/s, air exchange can improve from about 1.2^hr-1^ with windows closed to 4.0^hr-1^ with 2 windows open (Mutch 1985).

Another study observed 0-0.5 air exchanges per hour with windows and doors closed, 9 to 15 air exchanges per hour with the window opened, and about 40 air exchanges per hour with both windows and window-doors opened (Heidt 1981).

Other studies in homes have found 0 to 0.1 air changes per hour with low window use, 0.1 to 0.5 air changes per hour with average window use, and 0.5 to 0.8 air changes per hour with high window use (Dubrul 1988).

Another study of two homes found baseline air exchange rates of 0.4 hr^-1^, which increased by 0.1 to 2.8 hr^-1^ in one home and 0.5 to 1.7 hr^-1^ when windows were opened (Howard-Reed 2002).

Dick and colleagues (1951) estimated that typical households experienced 2 to 3 air exchanges per hour from opening windows. Air exchange rates also depend on wind speed (Dick 1951). With 5 mph wind, and 4 vents open, 2.8 air exchanges per hour were observed (Dick 1951).

For some perspective, it should be noted that in Germany, required ventilation of 40 m^3^/h per person for living spaces corresponds with about 0.5 to 0.6 exchanges per hour (Heidt 1981).

In addition to window-opening, ventilation will be positively correlated with temperature because in non-air conditioned environments, fans are used more frequently at higher temperatures (Haldi 2008; Nicol 1999; Rijal 2008; Rijal 2013; Honnekeri 2014), and doors are opened (Haldi 2008).

### Review of air conditioning

The final stage in this progression is to have central heating and air conditioning. Eventually, the windows might be left closed all the time, the central system controls ventilation, and the human is taken out of the loop. For instance, in one air-conditioned office building in Pakistan, occupants never opened the windows (Rijal 1988).

In U.S. residences, the presence of central air-conditioning was associated with a lower likelihood of window opening than a window-unit, which was itself lower than in homes without air conditioning (Johnson 2005).

In a hospital, window-opening and air exchange regressions suggest that rooms would experience 1.6 air exchanges per hour at 35 °F, and 2.5 air exchanges per hour at 60 °F (Baird 1969). Overall, winter conditions might see fresh air exchange rates of 0.5 per hour, compared with 2.0 exchanges per hour in summer (Baird 1969). Full air conditioning might provide 7.5 air changes per hour (Baird 1969).

In Korean homes, air conditioning was turned on at a temperature of 30 C, and turned off at a temperature of 26.9 C, in order to maintain the mean seasonal indoor temperature at 27.5 C (81.5 °F) (Bae 2009). In Japan, some people began to turn on air conditioners at 25 C, but most people did so between 28 and 30 C (Habara 2013). Another study in Japan found air conditioners turned on at an outdoor temperature of 30 C, in order to maintain the room at an average temperature of 29 C (Iwashita 1997).

Another study in Japan found that about half the residents had turned on the air conditioner at an indoor temperature of 27 C (Schweiker 2009). In one study of air-conditioned apartments in China, window opening frequency decreased above 24 C, and reached zero above 40 C (Shi 2016). In the United States, somewhat cooler temperatures were set. In 5 U.S. states, the median air-conditioning activation temperature was 74.9 °F (23.8 C) (Haiad 2004).

## Discussion

Our study found that the association of Covid-19 mortality with outdoor temperature shows two peaks. The global maximum of Covid-19 mortality occurs close to freezing, while a local minimum in Covid-19 mortality is observed close to room temperature. The local minimum of Covid-19 mortality at room temperature likely relates to improved ventilation from window opening and less indoor crowding when it is comfortable outside. Below freezing, all windows are closed, and further cooling increases stack ventilation (secondary to indoor-outdoor temperature differences) and thereby decreases Covid-19 mortality. Opening windows and other tools for improving indoor ventilation may decrease the spread of Covid-19.

It is important to understand that this analysis did not compare warm regions with cold regions. Rather, we compared warm weeks in one region with cold weeks in the same region. Because each area was compared with itself, regional variables such as population density, obesity and smoking prevalence, housing conditions, and willingness of the population to comply with mask wearing or restrictions on crowding were implicitly controlled for (to some degree).

Our study determined that the relationship between Covid-19 mortality and dew point was similar to that with temperature. Outdoor humidity has been positively correlated with window opening (Brundrett 1977, Jeong 2016, Andersen 2017) and indoor air change rates (Du 2012) in some studies, but other studies have found that the association of humidity with window opening was negative Johnson & Long 2005, Zhang 2012, Jones 2017), absent (Isaacs 2013), or multiphasic (Haldi 2009, Shi 2016).

Our study found that wind speed was negatively correlated with Covid-19 mortality. The association appeared somewhat smoother when effective wind speed was assumed to be zero in cold environments, to simulate the effect of window closure on the transmission of wind speed indoors. Indeed, wind speed has been positively correlated with indoor air change rates (Dick 1951).

The elevated Covid-19 mortality above room temperature might help to explain the higher mortality rates in the sunbelt of the United States in the summer of 2020. In addition, the elevated mortality rates observed in India in the spring of 2021 when the Delta variant began to predominate (Leffler 2021). For instance, the hottest months for Hyderabad, India are May, April, and March, due to the cooling that occurs from the rainy summer season (climate-data.org).

If the relationship between Covid-19 mortality and temperature had been a simple monotonic relationship, we might expect very high mortality in the coldest polar regions. The fact that mortality begins to decline at temperatures below freezing, presumably due to increased stack ventilation, might help to explain how many of the coldest Arctic regions, such as Iceland, Greenland, and some Scandinavian countries, have been able to avoid the highest levels of mortality.

We thought that window closure in areas with a high household share of air-conditioning might explain the second peak in Covid-19 mortality at temperatures above room temperature. However, this peak was present even in areas in which air conditioning was uncommon. There is some data that window opening may decrease a the very highest temperatures as occupants seek to protect themselves from the hot outdoor air (Herkel 2008). As we noted above, air conditioning can sometimes actually increase the ventilation rates (Baird 1969). Our study compared Covid-19 mortality at different times within a region, rather than comparing rates between regions. Nonetheless, we do note that many Asian countries in which air conditioning is popular have had low mortality rates during the pandemic (Leffler 2020).

These findings might have implications beyond Covid-19. It has been noted that influenza typically has a winter peak in temperate climates, but two annual peaks in the tropics (Ali 2021). This observation was recently attributed to a U-shaped relationship between transmission and absolute humidity (Ali 2021). Our findings and review would suggest that the association of environmental variables with indoor ventilation might be another fruitful avenue for investigation with respect to the incidence of respiratory disease.

The findings imply that in areas entering the winter period, rates of Covid-19 mortality could increase in areas expected to cool in the range between room temperature and freezing. Subtropical regions which cool only to room temperature during the winter are at less risk. Of course, regional outcomes will be influenced by both natural and vaccine-acquired immunity, non-pharmaceutical interventions, such as mask-wearing (Leffler 2020), and the flow of travelers from regions with different weather patterns.

## Data Availability

Data are available from the first author.

## References

Ali ST, Cowling BJ, Wong JY, Chen D, Shan S, Lau EH, He D, Tian L, Li Z, Wu P. Influenza seasonality and its environmental driving factors in mainland China and Hong Kong. Science of The Total Environment. 2021 Nov 17:151724.

American Housing Survey. 2019. Available from: https://www.census.gov/programs-surveys/ahs/data/interactive/ahstablecreator.html?s_areas=00006&s_year=2019&s_tablename=TABLE3&s_bygroup1=1&s_bygroup2=1&s_filtergroup1=1&s_filtergroup2=1 Accessed May 8, 2021.

Abergel T, Delmastro C. Is cooling the future of heating. International Energy Agency. December 13, 2020. Available from: https://www.iea.org/commentaries/is-cooling-the-future-of-heating Accessed Feb 7, 2021.

Alqahtani AS, Althobaity HM, Al Aboud D, Abdel-Moneim AS. Knowledge and attitudes of Saudi populations regarding seasonal influenza vaccination. Journal of Infection and Public Health. 2017 Nov 1;Nov(6):897–900.

Andersen RV. Occupant behaviour with regard to control of the indoor environment. Technical University of Denmark. 2009.

Arokiaraj MC. Correlation of Influenza Vaccination and the COVID-19 Severity. Available at SSRN 3572814. 2020 Apr 10.

Bae C, Chun C. Research on seasonal indoor thermal environment and residents’ control behavior of cooling and heating systems in Korea. Building and Environment. 2009 Nov 1; 44(11):2300–7.

Baird G. Air change and air transfer in a hospital ward unit. Building Science. 1969 Jan 1;Jan(3):113–24.

Brundrett GW. Ventilation: a behavioural approach. International Journal of Energy Research. 1977;1(4):289–98.

Brundrett, G.W., 1978, September. Window ventilation and human behaviour. In Proceedings of the first international indoor climate symposium in Copenhagen: indoor climate, effects on human comfort, performance and health in residential, commercial and light-industry buildings (pp. 317–330).

City Population. Major Agglomerations of the World. Available from: https://www.citypopulation.de/en/world/agglomerations/ Accessed November 25, 2020.

Climate Hyderabad (India). Climate-data.org. Available from: https://en.climate-data.org/asia/india/hyderabad/hyderabad-2801/ Accessed November 21, 2021.

Conner CC, Lucas RL. Thermostat related behavior and internal temperatures based on measured data in residences. No. PNL-7465. Pacific Northwest Lab., Richland, WA (USA), 1990.

Dash R, Agrawal A, Nagvekar V, Lele J, Di Pasquale A, Kolhapure S, Parikh R. Towards adult vaccination in India: a narrative literature review. Human Vaccines & Immunotherapeutics. 2020 Apr 2;Apr(4):991–1001.

Davies MG, Davies AD. The passive solar heated school in Wallasey. VII. Window opening behaviour and the microclimate of a classroom. International journal of energy research. 1987 Jul;11(3):315–26.

De Cian E, Pavanello F, Randazzo T, Mistry MN, Davide M. Households’ adaptation in a warming climate. Air conditioning and thermal insulation choices. Environmental Science & Policy. 2019 Oct 1; 100:136–57.

Dick JB, Thomas DA. Ventilation Research in Occupied Houses. Journal of the Institution of Heating and Ventilating Engineers. 1951; 19(194):306–26.

Dubrul C. Inhabitant behaviour with respect to ventilation—a summary report of IEA Annex VIII. Air Infiltration and Ventilation Centre. Berkshire, UK. 1988.

Erhorn H. Influence of meteorological conditions on inhabitants’ behaviour in dwellings with mechanical ventilation. Energy and Buildings. 1988;11:267–75.

Escobar LE, Molina-Cruz A, Barillas-Mury C. Correction for Escobar et al., BCG vaccine protection from severe coronavirus disease 2019 (COVID-19). PNAS. November 3, 2020 117 (44) 27741–27742; first published October 12, 2020; https://doi.org/10.1073/pnas.2019438117 Accessed November 8, 2020.

Escobar LE, Molina-Cruz A, Barillas-Mury C. BCG vaccine protection from severe coronavirus disease 2019 (COVID-19). Proceedings of the National Academy of Sciences. 2020 Jul 28;Jul(30):17720–6.

Garcia PJ, George PE, Romero C, Soto G, Carcamo C, Bayer AM. ‘The flu… is a little more complicated than a cold’: Knowledge, beliefs, and practices related to influenza and influenza vaccination among at-risk populations and health professionals in Peru. Vaccine. 2020 Oct 16.

Grech A, Yousif C. Lifestyle trends for heating and cooling in Maltese households. University of Malta. 2013. Available from: https://www.um.edu.mt/library/oar/bitstream/123456789/25474/1/No5_2013.pdf Accessed November 19, 2020.

Habara H, Yasue R, Shimoda Y. Survey on the occupant behavior relating to window and air conditioner operation in the residential buildings. In:13th Conference of International Building Performance Simulation Association. Chambery, France. 2013: 2007–2013.

Haiad C, Peterson J, Reeves P, Hirsch J. Programmable thermostats installed into residential buildings: Predicting energy savings using occupant behavior & simulation. Southern California Edison. 2004.

Haldi F, Robinson D. On the behaviour and adaptation of office occupants. Building and environment. 2008 Dec 1;Dec(12):2163–77.

Heidt FD, Haberda F, Trepte L. Impact of air infiltration and ventilation on energy losses of buildings. In: de Oliveira Fernandes E (ed.). Building Energy Management. Oxford: Pergamon Press, 1981: 201–214.

Herkel S, Knapp U, Pfafferott J. Towards a model of user behaviour regarding the manual control of windows in office buildings. Building and Environment. 2008 Apr 1;Apr(4):588–600.

Honnekeri A, Pigman MC, Zhang H, Arens E, Fountain M, Zhai Y, Dutton S. Use of adaptive actions and thermal comfort in a naturally ventilated office. UC Berkeley. 2014. Available from: https://escholarship.org/uc/item/37r4w5zs Accessed November 15, 2020.

Howard-Reed C, Wallace LA, Ott WR. The effect of opening windows on air change rates in two homes. Journal of the Air & Waste Management Association. 2002 Feb 1;Feb(2):147–59.

Huang Z, Huang J, Gu Q, Du P, Liang H, Dong Q. Optimal temperature zone for the dispersal of COVID-19. Science of The Total Environment. 2020 Sep 20;736:139487.

International Energy Agency. The Future of Cooling. Opportunities for energy-efficient air conditioning. Technology report — May 2018. Available from: https://www.iea.org/reports/the-future-of-cooling Accessed November 20, 2020.

International Energy Agency. The Future of Cooling in Southeast Asia. October 2019. Available from: https://www.iea.org/reports/the-future-of-cooling-in-southeast-asia Accessed November 22, 2020.

Iwashita G, Akasaka H. The effects of human behavior on natural ventilation rate and indoor air environment in summer—a field study in southern Japan. Energy and Buildings. 1997 Jan 1;Jan(3):195–205.

Japan Refrigeration and Air Conditioning Industry Association. World Air Conditioner Demand by Region. June 2019. Available from: https://www.jraia.or.jp/english/World_AC_Demand.pdf Accessed November 22, 2020.

Jeong B, Jeong JW, Park JS. Occupant behavior regarding the manual control of windows in residential buildings. Energy and buildings. 2016 Sep 1;127:206–16.

Johns Hopkins University. COVID-19 Data Repository by the Center for Systems Science and Engineering (CSSE) at Johns Hopkins University. Available from: https://github.com/CSSEGISandData/COVID-19 Accessed October 3, 2021.

Johnson T, Long T. Determining the frequency of open windows in residences: a pilot study in Durham, North Carolina during varying temperature conditions. Journal of Exposure Science & Environmental Epidemiology. 2005 Jul;15(4):329–49.

Kane T, Firth SK, Allinson D, Irvine K, Lomas KJ. Does the age of the residents influence occupant heating practice in UK domestic buildings. In: East Midlands Universities Association 2010 Conference-Perspectives in Society: Health, Culture, and the Environment, University of Leicester. 2010. Available from: http://mmmm.lboro.ac.uk/doc/EMUA%20conference%20paper.pdf Accessed November 15, 2020.

Karlinsky A, Kobak D. The World Mortality Dataset: Tracking excess mortality across countries during the COVID-19 pandemic. eLife 2021;10:e69336. Available from: https://elifesciences.org/articles/69336 Accessed July 25, 2021.

Khalfallah E, Missaoui R, El Khamlichi S, Ben Hassine H. Energy-Efficient Air Conditioning: A Case Study of the Maghreb. World Bank; 2016 Apr. Available from: http://documents1.worldbank.org/curated/en/754361472471984998/pdf/105360-REVISED-PUBLIC-MENA-Digital-Print-English-sep-2016.pdf Accessed November 20, 2020.

Kobak D, Karlinsky A. World Mortality Dataset. Excess Mortaliity. Available from: https://raw.githubusercontent.com/dkobak/excess-mortality/main/excess-mortality.csv Accessed October 12, 2021.

Korsavi SS, Montazami A, Brusey J. Developing a design framework to facilitate adaptive behaviours. Energy and Buildings. 2018 Nov 15;179:360–73.

Lamb B, Westberg H, Bryant P, Dean J, Mullins S. Air infiltration rates in pre-and post-weatherized houses. Journal of the Air Pollution Control Association. 1985 May 1;May(5):545–51.

Leffler CT, Ing E, Lykins JD, Hogan MC, McKeown CA, Grzybowski A. Association of Country-wide Coronavirus Mortality with Demographics, Testing, Lockdowns, and Public Wearing of Masks. Am J Trop Med Hyg. 2020 Dec;103(6):2400–2411.

Leffler CT, Lykins JD, Yang E. Preliminary analysis of excess mortality in india during the covid-19 pandemic (Update Sep. 26, 2021). medRxiv. 2021. Available from: https://www.medrxiv.org/content/10.1101/2021.08.04.21261604v2 Accessed November 21, 2021.

Li N, Li J, Fan R, Jia H. Probability of occupant operation of windows during transition seasons in office buildings. Renewable Energy. 2015 Jan 1;73:84–91.

Lin B, Wang Z, Liu Y, Zhu Y, Ouyang Q. Investigation of winter indoor thermal environment and heating demand of urban residential buildings in China’s hot summer–Cold winter climate region. Building and Environment. 2016 May 15;101:9–18.

Macroepidemiology of Influenza Vaccination (MIV) Study Group. The macroepidemiology of influenza vaccination in 56 countries, 1997–2003. Vaccine. 2005 Oct 25;Oct(44):5133–43.

Meyer D, Shearer MP, Chih YC, Hsu YC, Lin YC, Nuzzo JB. Taiwan’s annual seasonal influenza mass vaccination program—lessons for pandemic planning. American journal of public health. 2018 Sep;108(S3):S188–93.

Mutch RA. Heat losses due to window opening by occupants. In: CLIMA 2000 World Congress on Heating, Ventilating and Air-conditioning. Copenhagen. 1985; 2: Building design and performance: 25-32.

National Centers for Environmental Information. National Oceanic and Atmospheric Administration. Global Surface Summary of the Day. Available from: https://www.ncei.noaa.gov/access/search/data-search/global-summary-of-the-day Accessed October 20, 2021.

Nevius MJ, Pigg S. Programmable thermostats that go berserk? Taking a social perspective on space heating in Wisconsin. Energy Center of Wisconsin and Department of Sociology, University of Wisconsin. 2000:233–44.

Nicol JF, Raja IA, Allaudin A, Jamy GN. Climatic variations in comfortable temperatures: the Pakistan projects. Energy and buildings. 1999 Aug 1;Aug(3):261–79.

Oreszczyn T, Hong SH, Ridley I, Wilkinson P, Warm Front Study Group. Determinants of winter indoor temperatures in low income households in England. Energy and Buildings. 2006 Mar 1;Mar(3):245–52.

Pallot AC. Window opening in an office building. Year Book of the Heating and Ventilating Industry. 1962/3: 4–22.

Raja IA, Nicol JF, McCartney KJ, Humphreys MA. Thermal comfort: use of controls in naturally ventilated buildings. Energy and buildings. 2001 Feb 1;Feb(3):235–44.

Residential Energy Consumption Survey. 2009. Air Conditioning in Homes in Midwest Region, Divisions, and States, 2009 Million Housing Units. Available from: https://www.eia.gov/consumption/residential/data/2009/index.php?view=characteristics#ac Accessed May 8, 2021.

Rijal HB, Tuohy P, Humphreys MA, Nicol JF, Samuel A, Raja IA, Clarke J. Development of adaptive algorithms for the operation of windows, fans, and doors to predict thermal comfort and energy use in Pakistani buildings. American Society of Heating Refrigerating and Air Conditioning Engineers (ASHRAE) Transactions. 2008;114(2):555–73.

Rijal HB. Field investigation of comfort temperature and adaptive model in Japanese houses. PLEA 2013. 2013. Munich, Germany.

Rijal HB, Humphreys MA, Nicol JF. Development of a window opening algorithm based on adaptive thermal comfort to predict occupant behavior in Japanese dwellings. Japan Architectural Review. 2018 Jul;1(3):310–21.

Sahak MN, Arifi F, Hammond AA, Laurenson-Schafer HJ, Saeedzai SA, Safi H, Abubakar A, Elkholy A, Rasooly MH, Ikram AN. The 2018-19 influenza season in Afghanistan: Epidemiology and Virology. Available from: https://www.researchsquare.com/article/rs-12258/v1 November 12, 2020.

Santamouris M, Paravantis JA, Founda D, Kolokotsa D, Michalakakou P, Papadopoulos AM, Kontoulis N, Tzavali A, Stigka EK, Ioannidis Z, Mehilli A. Financial crisis and energy consumption: A household survey in Greece. Energy and Buildings. 2013 Oct 1;65:477– 87.

Schweiker M, Shukuya M. Comparison of theoretical and statistical models of airconditioning-unit usage behaviour in a residential setting under Japanese climatic conditions. Building and Environment. 2009 Oct 1;Oct(10):2137–49.

Shi S, Zhao B. Occupants’ interactions with windows in 8 residential apartments in Beijing and Nanjing, China. InBuilding Simulation 2016 Apr 1 (Vol. 9, No. 2, pp. 221–231). Tsinghua University Press.

Shipworth M, Firth SK, Gentry MI, Wright AJ, Shipworth DT, Lomas KJ. Central heating thermostat settings and timing: building demographics. Building Research & Information. 2010 Feb 1;Feb(1):50–69.

Sailor DJ, Pavlova AA. Air conditioning market saturation and long-term response of residential cooling energy demand to climate change. Energy. 2003 Jul 1;Jul(9):941–51.

Stazi F, Naspi F, D’Orazio M. Modelling window status in school classrooms. Results from a case study in Italy. Building and Environment. 2017 Jan 1;111:24–32.

Verdeil É. The contested energy future of Amman, Jordan: between promises of alternative energies and a nuclear venture. Urban Studies. 2014 May;51(7):1520–36.

Wallace LA, Emmerich SJ, Howard-Reed C. Continuous measurements of air change rates in an occupied house for 1 year: the effect of temperature, wind, fans, and windows. Journal of Exposure Science & Environmental Epidemiology. 2002 Jul;12(4):296–306.

Wehenkel C. Positive association between COVID-19 deaths and influenza vaccination rates in elderly people worldwide. PeerJ. 2020 Oct 1;8:e10112.

Weihl J. Monitored residential ventilation behavior: a seasonal analysis. Proceedings from the ACEEE 1986;7:7.281-7.295.

Weihl JS, Gladhart PM. Occupant behavior and successful energy conservation: Findings and implications of behavioral monitoring. InProceedings of the 1990 ACEEE Summer Study on Energy Efficiency in Buildings 1990 (Vol. 2, pp. 171–180).

Wikimedia Corp. List of cities by average temperature. Available from: https://en.wikipedia.org/wiki/List_of_cities_by_average_temperature Accessed November 22, 2020.

World Health Organization. BCG Immunization coverage estimates by country. Available from: https://apps.who.int/gho/data/view.main.80500?lang=en Accessed November 9, 2020.

Woolf SH, Chapman DA, Sabo RT, Zimmerman EB. Excess deaths from COVID-19 and other causes in the US, March 1, 2020, to January 2, 2021. JAMA. 2021 Apr 2.

Wu Y, Jing W, Liu J, Ma Q, Yuan J, Wang Y, Du M, Liu M. Effects of temperature and humidity on the daily new cases and new deaths of COVID-19 in 166 countries. Science of the Total Environment. 2020 Aug 10;729:139051.

Xie J, Zhu Y. Association between ambient temperature and COVID-19 infection in 122 cities from China. Science of the Total Environment. 2020 Jul 1;724:138201.

Zwerling A, Behr MA, Verma A, Brewer TF, Menzies D, Pai M. The BCG World Atlas: a database of global BCG vaccination policies and practices. PLoS Med. 2011; 8(3):e1001012. Available from: http://www.bcgatlas.org/index.php Accessed November 9, 2020.

